# Estimation of R0 for the spread of SARS-CoV-2 in Germany from Excess Mortality

**DOI:** 10.1101/2021.11.14.21266295

**Authors:** Juan Pablo Prada, Luca Estelle Maag, Laura Siegmund, Elena Bencurova, Liang Chunguang, Eleni Koutsilieri, Thomas Dandekar, Carsten Scheller

**Affiliations:** Department of Bioinformatics, Biocenter, Am Hubland, University of Würzburg, 97074 Würzburg, Germany; Institute of Virology and Immunobiology, University of Würzburg, Versbacher Str. 7, 97078 Würzburg, Germany

## Abstract

For SARS-CoV-2, R0 calculations report usually 2-3, biased by PCR testing increases. Covid-19-induced excess mortality is less biased.

We used data from Robert Koch Institute on Covid incidence, deaths, and PCR tests and excess mortality to determine early, policy-free R0 estimates with a serial interval of 4.7 days.

The PCR-based R0 value was 2.56 (95% CI 2.52-2.60) for Covid-19 cases and 2.03 (95%CI 1.96-2.10) for Covid-19-related deaths. As the number of PCR tests increased, R0 values were corrected accordingly, yielding 1.86 for Covid-19 cases and 1.47 for Covid-19 deaths, excess deaths were 1.34 (95% CI 1.32-1.37).

R0 is much lower than previously thought. This fits the observed seasonal pattern of infection across Europe in 2020-2021, including emergence of more contagious escape variants such as delta.

**One-Sentence Summary:** Excess mortality reveals infection speed in Covid-19 is surprisingly low with seasonal infection patterns and escape variants.

## Introduction

The basic replication number (R0) of a virus describes the average number of secondary infections caused by an infected individual in an immunologically still naive population ^1^. R0 is a key factor in predicting the spread of a virus in a population. It is also used to estimate the proportion of individuals required in a population to achieve herd immunity ^2^. In addition, the magnitude of R0 can also be used to predict whether a respiratory virus in temperate climates will develop a seasonal pattern of infection (as observed with influenza viruses and endemic coronaviruses) rather than continuous transmission throughout the year ^3^.

R0 is influenced not only by intrinsic characteristics of the pathogen, such as its infectivity and mode of transmission, but also by characteristics of the population under study. For respiratory viruses, there are several such extrinsic characteristics that have a significant impact on the probability of transmission and thus on R0: The density of a population, the number of persons living in a household and their average vulnerability to infections, other social factors that affect the number of close contacts between infected and uninfected persons (e.g., use of public transportation, work laws when ill, etc.), and also the climate of the area in which the population is located ^4^.

Based on data from 425 confirmed cases in Wuhan, R0 of SARS-CoV-2 was estimated to be 2.2 ^5^. Another report estimating R0 based on case reports in Wuhan yielded a higher R0 of 5.7 ^6^. This wide range of values is also reflected in a number of other analyses in which R0 was determined to be between 1.95 (WHO estimate) and 6.49 (all reviewed in ^7^). The German RKI assumes an R0 in the range of 2.8-3.8 ^8^ based on systematic reviews ^9,10,11^.

All these estimations of R0 have in common that they are based on incidences of SARS-CoV-2 infections detected by PCR. These estimations are therefore not only dependent on the characteristics of the population under study, but also on testing strategies (e.g. representative sampling, symptom-based testing, contact-based testing of index patients, etc.) as well as rapidly increasing numbers of available and performed tests during the early weeks of the pandemic (at least, if no mathematical corrections for this increase were performed).

Because SARS-CoV-2 infections have led to excess mortality in many countries worldwide ^12^, the increase in excess mortality can be used as a surrogate for SARS-CoV-2 infections in order to calculate R0 independent of testing strategies and testing capacity. Here, we determined R0 for SARS-CoV-2 infections in Germany during the early phase of the epidemic in February and March 2020 based on Covid-19-associated excess mortality. For comparison, we also calculated R0 from incidence data of SARS-CoV-2 infections corrected by the increase in test capacities, as well as R0 from incidence data of PCR-confirmed Covid-19-related deaths.

## Methods

### Databases

The number of Covid-19 cases, Covid-19-related deaths and SARS-CoV-2-PCR-tests was accessed from the RKI website ^13^. The definition of “Covid-19 case” used here is that of the RKI, which does not use the date of receipt of a positive PCR sample, but rather the date of illness, which in some cases is several days earlier ^13^. Excess mortality was calculated from data of the Federal Statistical Office ^14^. Mobility data was taken from the Apple™ website ^15^. All used datasets can be downloaded as excel file from the supplementary material S1.

### Calculation of excess mortality

To calculate excess mortality per calendar week, the number of weekly deaths ^14^ in 2020 was subtracted from the mean of weekly deaths in 2016-2019. For calculation of “adjusted excess deaths”, the excess mortality in calendar week 10 was tared to 0 in all age groups and the values of the following calendar weeks were adjusted accordingly.

### Calculation of R0

R0 was determined using the R package from Obadia et al. ^16^ in R version 3.60. We selected the exponential growth method of the package for calculation of R0. The mean serial interval (average time between successive infection cases) was simulated following a gamma distribution with mean equal to 4.7 (±SD 2.9)^17^. Weekly incidence values of excess mortality were converted to simulated daily incidence values using a gamma distribution. The R-script can be downloaded from the supplementary material S2.

## Results

### Determination of the time period that can be used for the calculation of R0

The governments of Germany and its states have taken several measures to contain the SARS-CoV-2 epidemic in Germany in early 2020, including canceling mass events (implemented March 9), closing schools (implemented March 16), closing stores (except grocery stores and pharmacies) and implementing social distancing rules prohibiting personal contact outside the family (implemented March 23) (Fig. 1A). All of these measures, as well as widespread media coverage of the SARS-CoV-2 epidemic in Germany, likely had an impact on the spread of SARS-CoV-2. Therefore, to estimate the value of R0 in Germany, it is imperative to include only data from time points that either predate the implementation of these measures or from time points when these measures could not yet have had an impact on the observed parameter used to calculate R0. As shown in Fig. 1A, people in Germany started to reduce their mobility from March 12, i.e., even a few days earlier than social distancing was officially introduced. Because the incubation period between SARS-CoV-2 infection and the onset of Covid-19 symptoms is on average 5-6 days ^8^, behavioral changes can lead to an impact on the number of disease cases no earlier than 5-6 days later. For our R0 calculations, we were therefore able to use incidence data of Covid-19 disease cases up to and including March 15 (calendar week 12) without risking that behavioral changes may have had an impact on the R value determined (Fig. 2C).

**Fig. 1:**
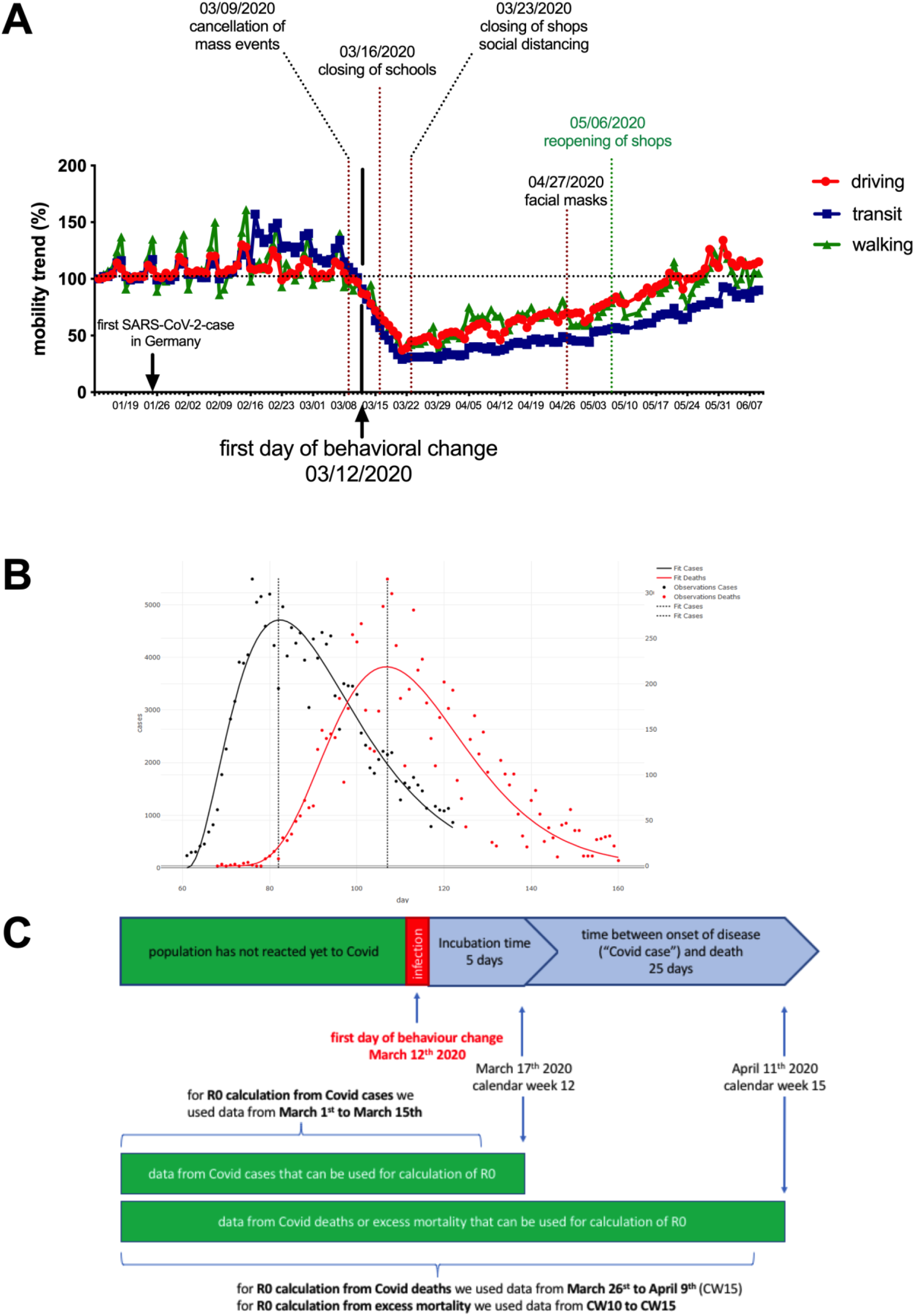
Identifying SRAS-CoV-2 datasets unaffected by policies or behavioral changes for estimating R0. **A:** Mobility data (driving) for Germany, provided by Apple™ Inc. ^15^ (driving: red; transit: blue; walking: green). The first change in mobility trends is observed for March 13. **B:** Data provided by RKI for Covid-cases (black) and Covid-19-related deaths (red) were fitted by gamma distribution. The maxima of the two curves are 25 days apart. **C**: Graphical representation of the date up to which data from Covid-19 disease cases or Covid-19 death cases can be used to determine R0 without affecting the outcome through policy actions or societal responses.

**Fig. 2:**
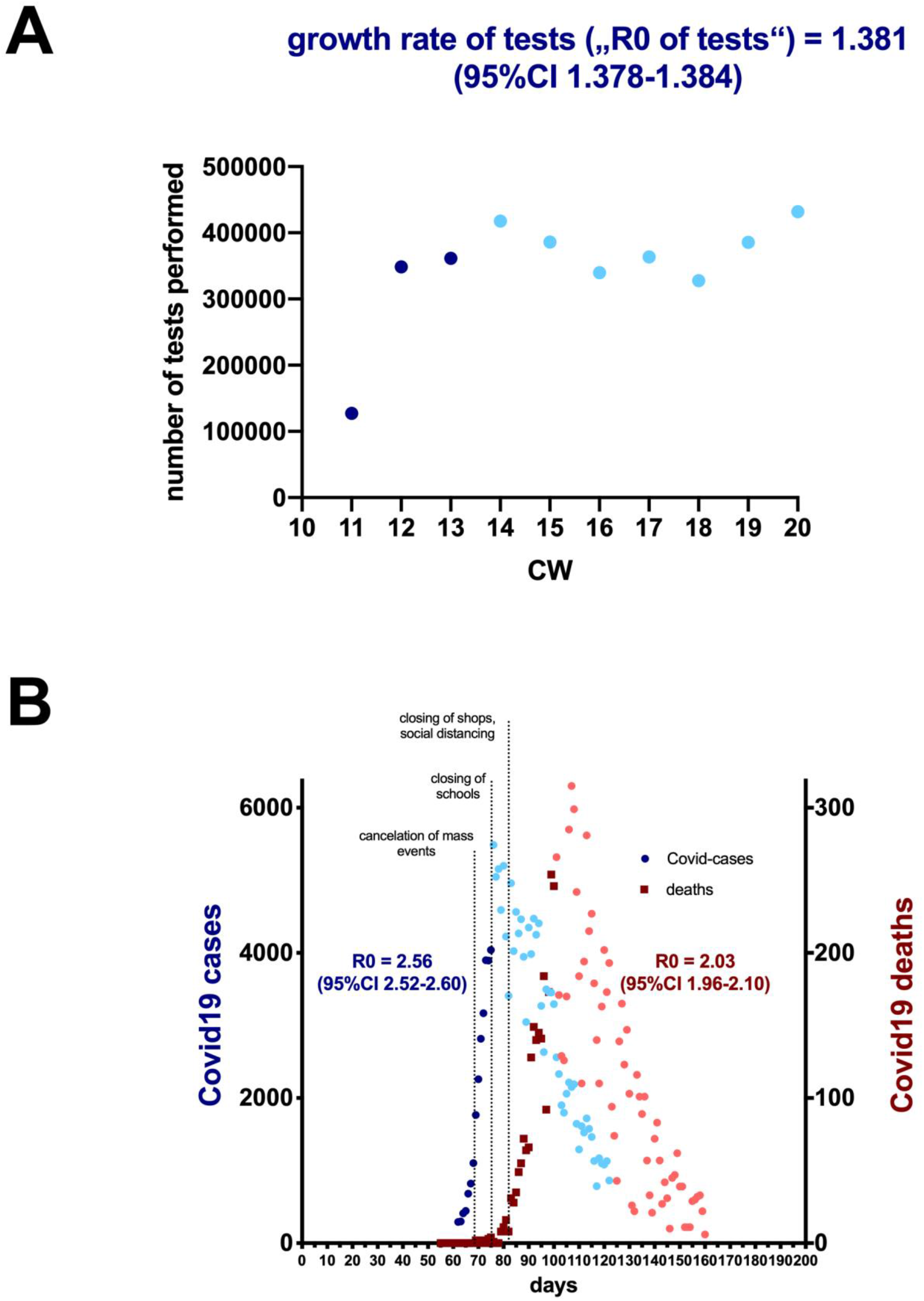
Calculation of R0 from Covid-19 disease incidence numbers and Covid-19 related deaths. **A:** Data reported by the RKI for the number of performed SARA-CoV-2 PCR-tests. **B:** Data reported by the RKI for Covid-19-cases (blue symbols, left y-axis) and Covid-19 related deaths (red symbols, right y-axis). A, B: Date were fitted to an exponential growth curve with a serial interval of 4.7 (±SD 2.9) to calculate R0. Dotted lines in B represent the dates for political interventions (03/09/2020 cancellation of mass events, 03/16/2020 closing of schools, 03/23/2020 closing of shops and social distancing). Dark blue and dark red symbols represent data points that were considered for the determination of R0, light blue and light red symbols represent later data points that were not considered for the calculation.

The German Robert Koch Institute (RKI) provides different epidemiological datasets that can be used for calculations of R0 of SARS-CoV-2 such as daily numbers of detected cases and daily numbers of CoViD-19-related deaths. We fitted the data to a gamma distribution and determined the difference between the peaks of the curves. The mean time between the occurrence of Covid-19 disease cases and Covid-19-related deaths was 25 days (Fig. 1B). Covid-19-related deaths can therefore be used for the determination of R0 at significantly later time points than the occurrence of Covid-19 disease cases without risking of compromising the R0 value by behavioral changes. Therefore, to determine R0 from reported Covid-19 deaths (as well as from Covid-19-related excess mortality), we used records up to and including April 11 (calendar week 15) (Fig. 2C).

### Calculation of R0 from incidence data of Covid-19 disease cases and Covid-19 deaths

In the initial phase of the pandemic, testing capacities were significantly smaller than the actual number of infections. The steep rise in the number of reported cases during this period might therefore also due in significant part to the sharp increase in the number of SARS-CoV-2 PCR test performed. For our calculations of R0, we use incidence data up to and including calendar week 12 for Covid-19 disease cases and up to and including calendar week 15 for Covid-19 deaths. While data for the number of tests performed are not available for the period before calendar week 11, the RKI provides at least the number of tests performed from week 11 onwards ^13^. As depicted in fig. 2A, a significant increase in the number of tests performed can be observed in the calendar weeks 11 to 13. To determine what impact this increase in testing numbers had on reported Covid-19 incidences, we determined the growth rate of testing during this period. The growth rate of testing yields an “R0 of tests” of 1.38 (Fig. 2A), meaning that even if the number of infections remained constant during this period of time, an apparent increase of 1.38 in R0 would be observed. It follows that R0 values from incidence figures must be corrected by this factor.

From the raw incidence data, we obtain an R0 of 2.56 for Covid-19 disease cases and an R0 of 2.03 for Covid-19 death cases (Fig. 2B). However, these values must still be corrected for the growth rate of testing (R0_corrected_ = R0_uncorrected_/”R0 of tests”), resulting in a corrected R0 of 1.86 for Covid-19 disease cases and an R0 of 1.47 for Covid-19-death cases. The R0 value derived from deaths is slightly lower than the R0 value determined from Covid-19 incidence values. This may be due to the fact that in the initial phase of the pandemic, severely ill cases (and thus individuals at higher risk of death) were preferentially tested, while milder and asymptomatic cases were increasingly included in testing as the testing capacity expanded. Such a change in testing strategy inevitably introduces a bias toward higher R0 values when calculated from Covid-19 incidence data compared with Covid-19 death data.

However, even if we correct the incidence values for test capacity dynamics, this way of determining R0 still remains subject to many uncertainties: First, the exact numbers of tests performed in the first weeks of the pandemic were not collected for Germany, so that an accurate estimate of the dynamics of testing capacity is not possible. Second, the incidence data do not come from representative samples in the general population, but mainly from symptomatic patients and persons with whom they came into contact. Therefore, this dataset contains a disproportionate number of infections from nursing homes and hospitals, where symptomatic infections are overrepresented and where transmission probabilities are most likely different from what would be expected in the general population. Therefore, the R0 values calculated above are unlikely to be representative of the spread of the virus in the general population.

### Calculation of R0 from Excess mortality

To address this problem, we also determined R0 based on excess mortality data in Germany during early 2020. The Federal Statistical Office of Germany lists all deaths that occur in Germany, regardless of their cause ^14^. Because SARS-CoV-2 infection has led to increased excess mortality in many countries, these data can be used as surrogate markers for the spread of SARS-CoV-2 infections. And because excess mortality is independent of the number or strategy of SARS-CoV-2 testing, it provides a representative picture for the spread of infections in the general population.

Figure 3A shows the incidence of deaths with confirmed SARS-CoV-2 infection. The data set shown here is the same as that in Figure 2B, but this time as weekly incidence and subdivided into different age groups. The (uncorrected) R0 value here is 1.95, similar to 2.03 from figure 2B. From this figure, it can be seen that the peak of Covid-19 related mortality is between calendar week 10 and 20. Figure 3B shows excess mortality (in relation to average weekly deaths in 2016-2019) in the different age groups, and one can see a parallel trend to the confirmed Covid-related deaths between calendar weeks 10 and 20. Based on the respective values of calendar week 10, from which an increase in excess mortality is observed in all Covid-19 relevant age groups, we plotted the change in all values in Figure 3C. From this adjusted excess mortality, we obtained an R0 of 1.34 (95% CI 1.32-1.37) for the sum of all age groups for the spread of SARS-CoV-2 in the general population in Germany.

**Fig. 3.**
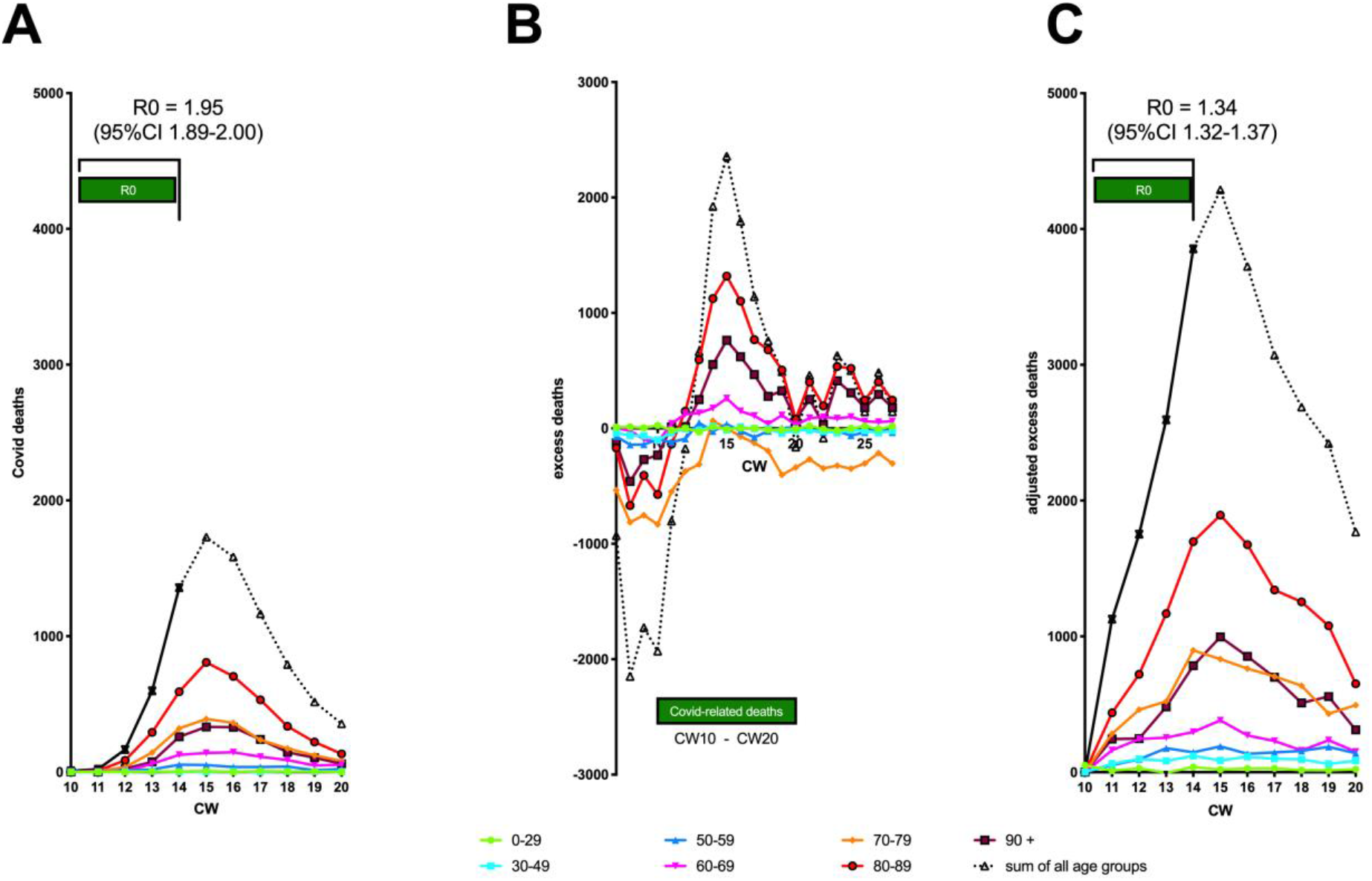
Calculation of R0 from excess mortality. **A:** Covid-19 related deaths as weekly incidence in different age groups. **B**: Excess deaths in 2020 in different age groups based on comparison with average weekly deaths in 2016-2019. **C**: Excess deaths presented in B, but adjusted to 0 for week 10 in each age group, so that relative changes related to Covid-19 become better visible.

### Influence of Influenza-related excess deaths on Covid-19 related excess deaths

Due to the lack of representative measurements, Covid-19 related excess mortality is the only infection parameter that is free from bias due to changes in testing strategy or testing numbers. However, excess mortality data are subject to other confounding factors that may have an impact on the calculation of R0: The Covid-19 pandemic reached Germany at a time when seasonal influenza activity in Germany was already subsiding (see Fig. 4C). Influenza-related excess mortality and Covid-19 related excess mortality are therefore superimposed in the total excess mortality data sets. If influenza mortality had been significantly elevated in 2020 compared with previous years (2016-2019), this could mask Covid-19 related effects, particularly if influenza mortality rates that were already falling again coincided with an incipient increase in Covid-19 mortality rates. However, a look at mortality in previous years shows that influenza-related mortality in Germany in 2020 was significantly lower compared with previous years because of the two exceptionally strong influenza years 2017 and 2018 (Fig. 4A). As a result, under-mortality was observed in Germany in calendar weeks 10-14 compared with previous years, rather than excess-mortality (Fig. 4B). Thus, if there was an effect of influenza-related deaths on the calculation of R0 for Covid-19 infections, it was one that resulted in an overestimate of R0 rather than an underestimate. Thus, the R0 calculated here for R0 of SARS-CoV-2 of 1.34 (95% CI 1.32-1.37) should be regarded as a maximum value, whereas the actual R0 of SARS-CoV-2 infections in Germany is likely to be even lower.

**Fig. 4:**
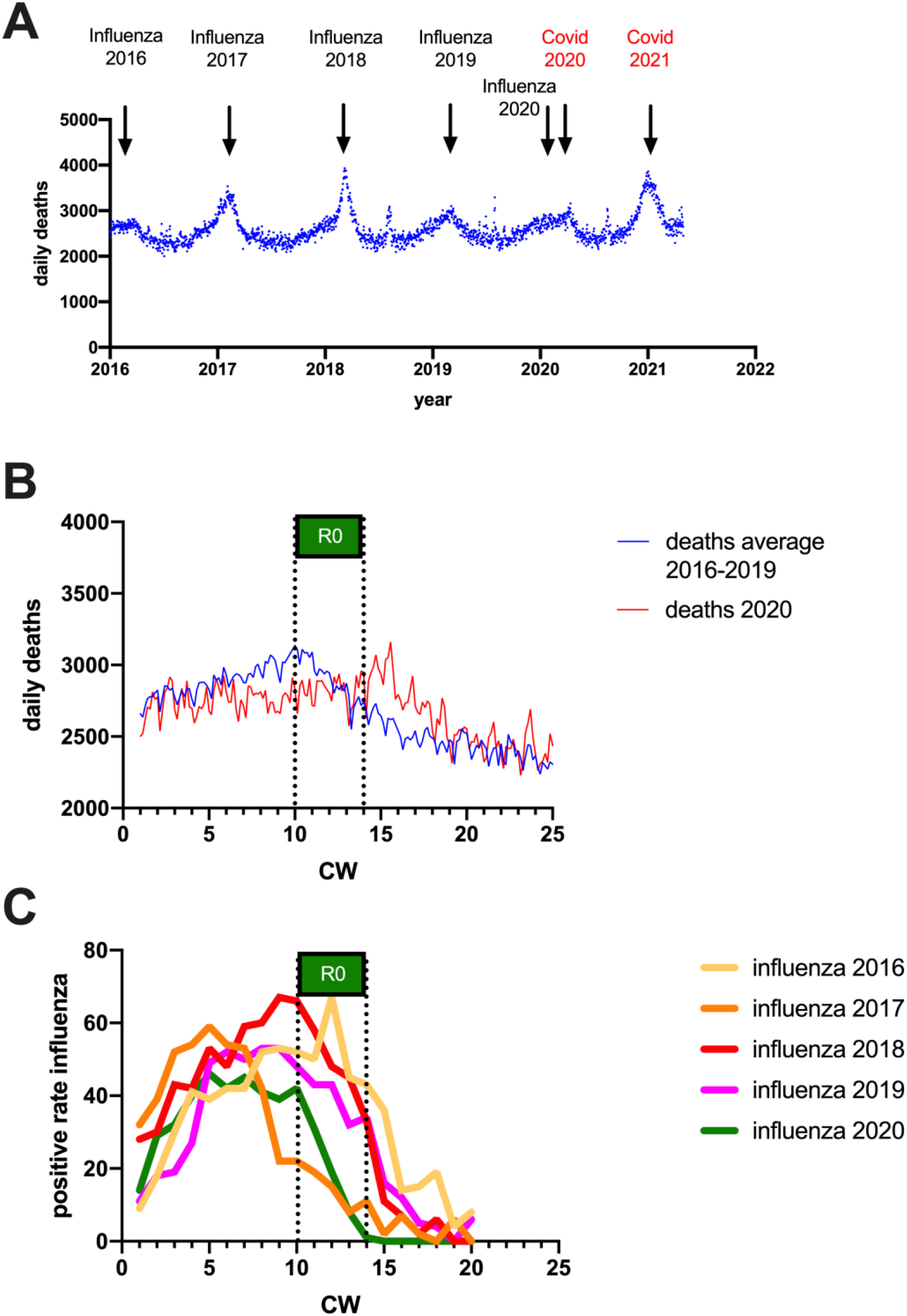
Comparison of Influenza-related and Covid-19 related mortality in Germany. **A:** Daily deaths in Germany from 2016 to 2021 (data taken from the Federal Statistical Office of Germany). **B:** Comparison of daily number of deaths in 2020 (red line) with average number of daily deaths in 2016-2019 (blue line). **C:** Positive rate for influenza infections in Germany for calendar weeks 1-20 from 2016-2020 (data taken from the RKI influenza survey).

## Discussion

The early SARS-CoV-2 infection spread in Germany with an R0 of 1.34 (95% CI 1.32-1.37). This value is much lower than what had been expected based on R0 determinations from the literature, where values between 2-3 became consensus ^7,8^. Although the German RKI has not published an R0 estimation for Germany, it provided daily estimations of R based on a four-day-period. These daily R values during the first two weeks of March 2020 were in the range of 2.2-3.2 ^18^. Based on the reporting data for positive PCR results from the “our world in data” database of Oxford University, an R0 of 3.37 was determined for Germany ^19^, but these calculations used PCR reporting data rather than data for Covid-19 disease cases and therefore are not fully comparable with the calculations from the RKI. The discrepancies between these high values and the rather low R0 estimates in our manuscript are primarily due to the fact that in these earlier estimations the R0 values were not corrected by a factor accounting for the substantial increase in test capacity during this period. If we use our uncorrected R0 estimate based on Covid-19 case numbers for comparison (R0 = 2.56, Fig. 2B), it is in the same order of magnitude as the values calculated by the RKI.

A high R0 of the order of 3 would likely have resulted in a lack of seasonal progression, as a seasonal effect was estimated to reduce R0 by only 40% based on observations in endemic coronaviruses ^3^. Accordingly, health authorities expected an unrestrained spread of the virus for Germany, whereupon a series of policy measures were adopted aiming to actively reduce the incidence of infection. In retrospect, however, a clearly seasonal occurrence is evident not only for Germany, but also for all other countries in temperate climates, in particular also for Sweden, where hardly any measures have been taken to contain the spread of SARS-CoV-2 in the general population ^12^.

The seasonal effect on R0 can be approximated as a sinusoidal pattern with a maximum in January as the coldest month in Germany and a minimum in July as the warmest month, with an approximate 40% reduction in July compared to January ^3^ (Fig. 5, dotted line). As our calculation of R0 was based on the infection situation in March 2020, it can be expected that the R0 value determined in this way is about 20% lower than the maximum value that would be reached in January if the pandemic would have reached Germany earlier. According to this model, R0 determined in March with a value of R0_March_ = 1.34 would reach its maximum in January with a value of R0_January_ = 1.68 and fall to a minimum of R0_July_ = 1.01 in July (Fig. 5A). Herd immunity is dependent on R0 (herd immunity = (1-1/R0)), and therefore also fluctuates seasonally. Thus, herd immunity against the original SARS-CoV-2 strain would oscillate between 40% in January and below 1% in July (Fig. 5B). This explains why the number of SARS-CoV-2 infections in summer 2020 not only declined again in “lockdown” countries such as Germany (and remained low in these countries even after the suspension of policy measures during the summer), but also why the same seasonal pattern was observed in countries with little or no countermeasures against SARS-CoV-2 ^12^.

**Fig. 5:**
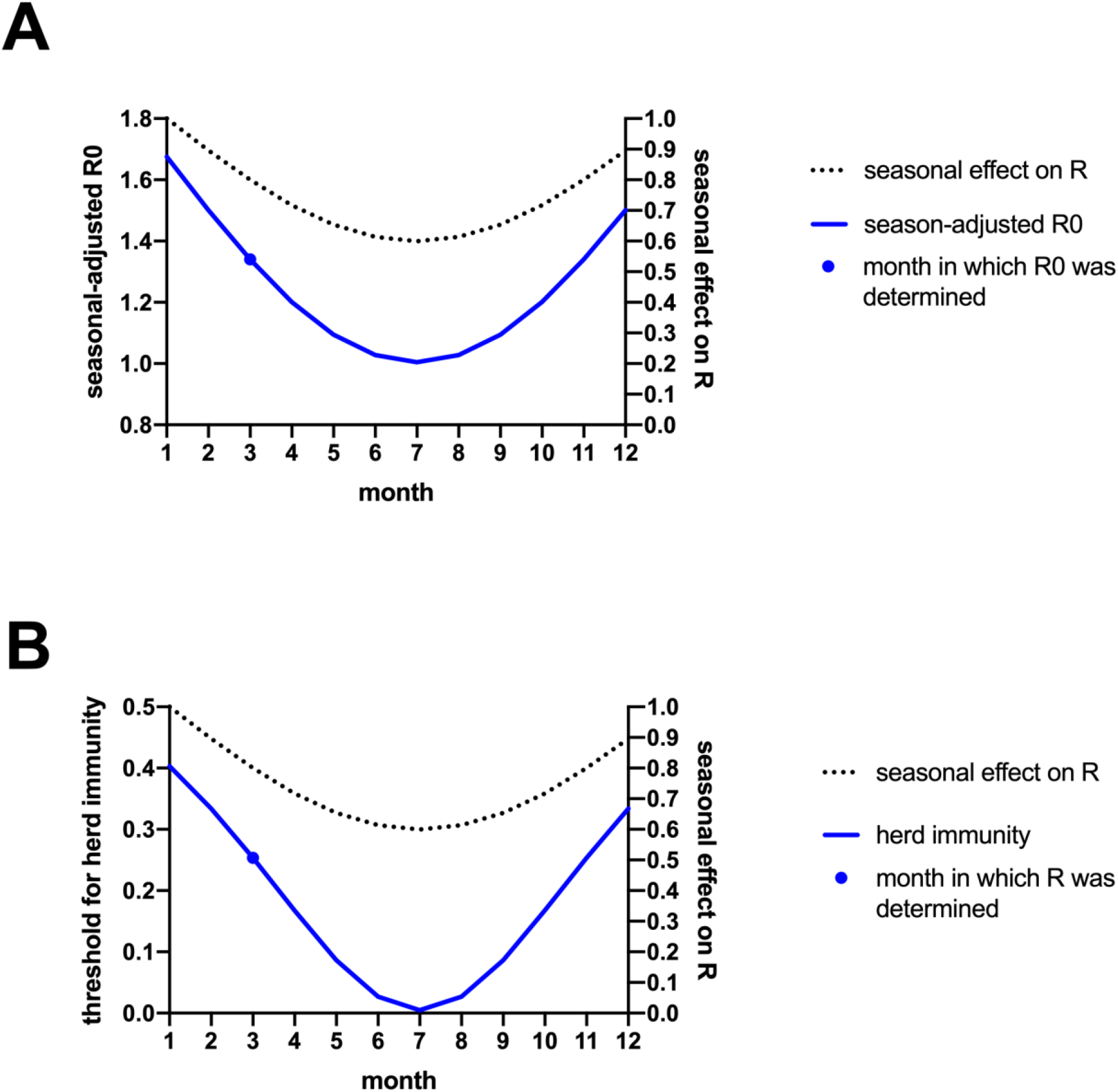
Seasonal influence on R0 and herd immunity. **A:** The seasonal effect on R0 can be assumed as a sine function with a maximum in January and a 40% lower minimum in July (dashed line, right y-axis). This translates into an oscillating R0 with a maximum in January (of R0_January_ = 1.68) and a minimum in July (R0_July_ = 1.01) (blue line, left y-axis). R0 calculations are based on the values calculated for March (R0_March_ = 1.34) (blue dot) **B:** Herd immunity similarly to R0 oscillates in a seasonal pattern.

With a seasonal increase in the threshold of herd immunity, the following winter (2020/2021) fueled again the spread of SARS-CoV-2 Germany, until infection numbers dropped again in spring 2021. At this time, about 3.8 million SARS-CoV-2 infections have been reported to the RKI, corresponding to 4.5% of the population (May 2021) ^20^ and a serological survey of blood donors revealed a serological positive rate of 14% in April 2021 ^21^, showing a substantial underestimation of SARS-CoV-2 infections from PCR-data alone. Together with the enrollment of the Covid-vaccine campaign, immunity in the German population has reached up to 40% by end of May 2021, and that coincided with the emergence of the delta variant, which can now be interpreted as an escape variant that overcame the 40% herd immunity restrictions of the original SARS-CoV-2 strains by higher contagiousness (and therefore also a higher R0). Since the population in Germany was no longer naïve towards SARS-CoV-2 during summer of 2021 when the delta variant began its expansion in Germany, it is not possible to determine an R0 for this variant. However, the RKI calculates daily R_e_ values based on a 7-day period for Germany, and the values for the delta variant reached 1.3 during July/August 2021^22^. With a seasonal variation of 40%, this value corresponds to a theoretic maximum in December with a R_e_ = 2.2, translating into a 55% threshold for winter herd immunity. Approximately 65% of the population in Germany has now been vaccinated against SARS-CoV-2 (by end of October 2021). Recent studies show that vaccine efficiency to protect from infection is comparatively rapidly decaying. A cohort study from September 2021 found that Covid vaccines are only 35% protective against infection^23^, and it is reasonable to assume that the vaccines will become even less effective as time after vaccination increases^28,29^. The proportion of the population in Germany already immune to SARS-CoV-2 infection can therefore only be estimated very roughly. To the approx. 20% who have already been infected, an estimated 20% immunity due to the vaccinations can be added (vaccination rate multiplied by effectiveness), so that we assume that the proportion of immunity towards SARS-CoV-2-infection in Germany is less than expected due to waning immunity. In spite of this, the protection by vaccination against severe infection and death by SARS-CoV-2 infection is well established to be high^30^. However, this is not the focus here but rather transmission of the virus taking also recent and longer time ago previously vaccinated people also into account, and thus the influence of all these different factors on the resulting R0.

The concept of herd immunity in respiratory pathogens such as coronaviruses does not imply permanent protection of the population against reemergence of these pathogens, since the immunity achieved may decrease over time, especially in asymptomatically infected patients ^24^. Instead, the achievement of herd immunity in respiratory viruses leads to a strong selection pressure for escape mutations (classical immune escape or increased contagiousness), which can then give rise to new waves of infection ^25^. For this reason, respiratory viruses such as influenza-or coronaviruses remain endemic, despite broad immunity, which will probably also be the case for SARS-CoV-2.

## Conclusion

Our study shows that the R0 value of SARS-CoV-2 can be calculated from excess mortality data. We also introduce here the concept of a seasonally adjusted R0 value, which should be reported as a range (R0_January_-R0_July_) rather than a static value. We determined an R0 value of 1.34 for infections in March 2020 (R0_March_ = 1.34), corresponding to a seasonal range of R0_January_ = 1.68 and a minimum in July (R0_July_ = 1.01). This rather low range of R0 values is much more consistent with observations of pandemic progression than many earlier and much higher estimates of the R0 value. The massive expansion of testing capacity in the early phase of the pandemic, combined with changes in testing strategy, was a major cause of the overestimation of the R0 value. Excess mortality can be determined independently of SARS-CoV-2 testing capacity in many countries, and therefore can be a valuable tool in future pandemics to provide reliable values for the rate of spread of an emerging pathogen in a population when representative samples of pathogen spread are not available.

## Data Availability

All data produced are available online at
https://www.rki.de/DE/Home/homepage_node.html

https://www.rki.de/DE/Home/homepage_node.html

## Acknowledgements

We thank the Land of Bavaria for funding. CL and TD specifically acknowledge the Land of Bavaria contribution to our DFG project 324392634-TRR 221/INF. For the preparation of this manuscript, we used the online version of DeepL translator ^26^ and Mendeley Reference Manager ^27^.

None.

## Supplementary material

S1 Database

S2 R Script

## Funding

We acknowledge Land of Bavaria contribution to DFG project 324392634-TRR 221/INF (TD) and DFG project 374031971-TRR 240/INF (TD).

## Author contributions

Conceptualization: CS

Methodology: JPP, TD, CS

Investigation: JPP, LEM, LS, EB, LC, EK, TD, CS

Visualization: JPP, LEM, LS, EB, LC,

Funding acquisition: TD, CS

Project administration: TD, CS

Supervision: EK, TD, CS

Writing – original draft: JPP, CS

Writing – review & editing: JPP, LEM, LS, EB, LC, EK, TD, CS

## Competing interests

Authors declare that they have no competing interests.

## Data and materials availability

All data are available in the main text or the supplementary materials, public data used are all noted and accessible.

## Supplementary Materials

Data sheet (excel)

R-script (txt) with README (txt)

## README

The script attached should simply be run on R, please simply adjust the path to the “datasheet_R0.xlsx” file.

**The datasheet excel file is given independent of this combined pdf as this is an excel file**.

This is the R session info:

**R version 3.6.0 (2019-04-26)**

**Platform: x86_64-w64-mingw32/x64 (64-bit)**

**Running under: Windows >= 8 x64 (build 9200)**

**Matrix products: default**

**locale:**

**[1] LC_COLLATE=German_Germany.1252 LC_CTYPE=German_Germany.1252**

**LC_MONETARY=German_Germany.1252 LC_NUMERIC=C LC_TIME=German_Germany.1252**

**attached base packages:**

**[1] stats graphics grDevices utils datasets methods base**

**other attached packages:**

**[1] R0_1.2-6 MASS_7.3-53.1 readxl_1.3.1**

**loaded via a namespace (and not attached):**

**[1] Rcpp_1.0.6 fansi_0.4.2 utf8_1.2.1 crayon_1.4.1 cellranger_1.1.0 lifecycle_1.0.0**

**magrittr_2.0.1 pillar_1.6.0 cli_2.5.0**

**[10] rlang_0.4.10 rstudioapi_0.13 vctrs_0.3.7 ellipsis_0.3.1 tools_3.6.0 yaml_2.2.1**

**compiler_3.6.0 pkgconfig_2.0.3 tibble_3.1.1**

~~~
R-Script
## script that accompany the supplemental material of the paper Estimation of R0 for the spread
of SARS-CoV-2 in Germany from Excess Mortality
## by Prada et. al.

## load the required packages, if not present these must be installed #########
require(readxl)
require(R0)

##### replace here the path where the file datasheet_R0 is #######################
path = “D:/Project-covid19/datasheet_R0.xlsx”

######################################### tests performed Fig. 2A
#############################################################
test_performed_per_week <-read_excel(path,sheet = “tests performed Fig. 2A”)[1:3,1:2]
##### re-structure the data to daily incidence
daily_incidences = matrix(0,ncol = ncol(test_performed_per_week),nrow =
nrow(test_performed_per_week)*7)
pos_rows = seq(1,nrow(test_performed_per_week)*7,7)
for (ic in 1:ncol(test_performed_per_week)) {
   for (ir in 1:nrow(test_performed_per_week)) {
      daily_incidences[pos_rows[ir]:(pos_rows[ir]+6),ic] = rmultinom(1, size =
as.numeric(test_performed_per_week[ir,ic]), prob = c(1/7,1/7,1/7,1/7,1/7,1/7,1/7))
   }
}
colnames(daily_incidences) = c(“day”,”number of tests”)
daily_incidences[,1] = 1:(nrow(test_performed_per_week)*7)
##### calculation of the RO value
GT.flu_w <-generation.time(“gamma”, c(4.7,2.9)) # daily time
R0 <-estimate.R(daily_incidences[,2], GT=GT.flu_w, methods=c(“EG”))
print(paste(“Tests performed Fig. 2A estimated R0 = “,R0$estimates$EG$R, “ CI
(“,R0$estimates$EG$conf.int[1],” -“,R0$estimates$EG$conf.int[2],”)”,sep=““))

######################################### Covid-cases and deaths Fig. 2B
#############################################################
covid_deaths_daily <-read_excel(path,sheet = “Covid-cases and deaths Fig. 2B”)[1:46,1:2]
covid_cases_daily <-read_excel(path,sheet = “Covid-cases and deaths Fig. 2B”)[8:21,c(1,3)]
##### calculation of the RO value
GT.flu_d <-generation.time(“gamma”, c(4.7,2.9)) # daily time
R0_d <-estimate.R(covid_deaths_daily$‘covid deaths’, GT=GT.flu_d, methods=c(“EG”))
print(paste(“Covid-deaths Fig. 2B estimated R0 = “,R0_d$estimates$EG$R, “ CI (“,R0_d$estimates$EG$conf.int[1],” -“,R0_d$estimates$EG$conf.int[2],”)”,sep=““))
R0_c <-estimate.R(covid_cases_daily$‘covid cases’, GT=GT.flu_d, methods=c(“EG”))
print(paste(“Covid-cases Fig. 2B estimated R0 = “,R0_c$estimates$EG$R, “ CI
(“,R0_c$estimates$EG$conf.int[1],” -“,R0_c$estimates$EG$conf.int[2],”)”,sep=““))

######################################### Covid-deaths Fig. 3A
#############################################################
ExcDeaths_per_week <-read_excel(path,sheet = “Covid-deaths Fig. 3A”)[1:4,1:9]
##### re-structure the data to daily incidence
daily_incidences = matrix(0,ncol = ncol(ExcDeaths_per_week),nrow =
nrow(ExcDeaths_per_week)*7)
pos_rows = seq(1,nrow(ExcDeaths_per_week)*7,7)
for (ic in 1:ncol(ExcDeaths_per_week)) {
   for (ir in 1:nrow(ExcDeaths_per_week)) {
     daily_incidences[pos_rows[ir]:(pos_rows[ir]+6),ic] = rmultinom(1, size =
as.numeric(ExcDeaths_per_week[ir,ic]), prob = c(1/7,1/7,1/7,1/7,1/7,1/7,1/7))
      }
}
colnames(daily_incidences) = colnames(ExcDeaths_per_week)
daily_incidences[,1] = 1:(nrow(ExcDeaths_per_week)*7)
##### calculation of the RO value
GT.flu_d <-generation.time(“gamma”, c(4.7,2.9)) # daily time
print(“Excess deaths Fig. 3A results”)
for (ind_col in 2:ncol(daily_incidences)) {
  final = floor(length(daily_incidences[,ind_col]))
  R0 <-estimate.R(daily_incidences[,ind_col], GT=GT.flu_d, methods=c(“EG”),begin =
1,end=final)
  print(paste(colnames(daily_incidences)[ind_col],” excess deaths estimated R0 =
“,R0$estimates$EG$R, “ CI (“,R0$estimates$EG$conf.int[1],” -
“,R0$estimates$EG$conf.int[2],”)”,sep=““))
}
######################################### Adjusted Excess deaths Fig. 3C
#############################################################
AdExcDeaths_per_week <-read_excel(path,sheet = “Adjusted Excess deaths Fig.
3C”)[1:4,1:9]
AdExcDeaths_per_week[AdExcDeaths_per_week<0] = 0 # excess death can be bellow 0 but
the incidence cannot
##### re-structure the data to daily incidence
daily_incidences = matrix(0,ncol = ncol(AdExcDeaths_per_week),nrow =
nrow(AdExcDeaths_per_week)*7)
pos_rows = seq(1,nrow(AdExcDeaths_per_week)*7,7)
for (ic in 1:ncol(AdExcDeaths_per_week)) {
   for (ir in 1:nrow(AdExcDeaths_per_week)) {
     daily_incidences[pos_rows[ir]:(pos_rows[ir]+6),ic] = rmultinom(1, size =
as.numeric(AdExcDeaths_per_week[ir,ic]), prob = c(1/7,1/7,1/7,1/7,1/7,1/7,1/7))
    }
}
colnames(daily_incidences) = colnames(AdExcDeaths_per_week)
daily_incidences[,1] = 1:(nrow(AdExcDeaths_per_week)*7)
##### calculation of the RO value
GT.flu_d <-generation.time(“gamma”, c(4.7,2.9)) # daily time
print(“Adjusted excess deaths Fig. 3B results”)
for (ind_col in 2:ncol(daily_incidences)) {
  R0 <-estimate.R(daily_incidences[,ind_col], GT=GT.flu_d, methods=c(“EG”))
  print(paste(colnames(daily_incidences)[ind_col],” adjusted excess deaths estimated R0 =
“,R0$estimates$EG$R, “ CI (“,R0$estimates$EG$conf.int[1],” -
“,R0$estimates$EG$conf.int[2],”)”,sep=““))
}
~~~

## Notes

### Competing Interest Statement

The authors have declared no competing interest.

### Funding Statement

This study was funded by Land of Bavaria as contribution to our DFG project 324392634-TRR 221/INF.

### Author Declarations

This study contains data from incidence of Covid-19 and data of excess death from Germany, all publicly available.

